# Aortic stenosis is independently associated with male gender, obesity renal failure, COPD, Caucasians and numerous inflammatory diseases in addition to known cardiovascular risk factors

**DOI:** 10.1101/2023.07.20.23292970

**Authors:** Mohammad Reza Movahed, Brandon Timmerman, Mehrtash Hashemzadeh

## Abstract

**Background:** Aortic valve stenosis is associated with age, rheumatic fever, and bicuspid aortic valve but its association with other comorbidities such as inflammatory disease and race is less known. The purpose of this study was to investigate any association between aortic stenosis and many comorbidities.

**Method:** We utilized the large Nationwide Inpatient Sample database to evaluate any association between aortic stenosis and risk factors. We performed uni- and multivariate analyses adjusting for comorbid conditions.

**Results:** Data were extracted from the first available database that used ICD-10 codes specifically coding for aortic stenosis alone, spanning from 2016 to 2020. Data included 112,982,565 patients. A total of 2,322,649 had aortic stenosis, with the remaining 110,659,916 patients serving as controls. We found a strong and independent significant association between aortic stenosis and coronary artery disease (OR: 2.11, CI 2.09 - 2.13, P < 0.001), smoking (OR: 1.08, CI 1.07 - 1.08, P < 0.001), diabetes mellitus (OR: 1.15, CI 1.14 - 1.16, P < 0.001), hypertension (OR: 1.41, CI 1.4 - 1.43, P < 0.001), hyperlipidemia (OR: 1.31, CI 1.3 - 1.32, P < 0.001), renal disease (OR: 1.3, CI 1.29 - 1.31, P < 0.001), chronic obstructive lung disease (COPD) (OR: 1.05, CI 1.04 - 1.05, P < 0.001), obesity (OR: 1.3, CI 1.29 -1.32, P < 0.001), rheumatoid arthritis (OR: 1.13, CI 1.11 - 1.15, P <0.001), scleroderma (OR: 1.93, CI 1.79 - 2.09, P <0.001), systemic connective tissue disease (OR: 1.24, CI 1.2 - 1.27, P <0.001), polyarteritis nodosa (OR: 1.5, CI 1.24 -1.81, P <0.001), and Raynauds syndrome (OR: 1.16, CI 1.09 - 1.24, P <0.001), in addition to known factors such as age, male gender and bicuspid aortic valve.

**Conclusion:** Using a very large database, we found many new associations for presence of aortic valve stenosis including race, renal disease, several inflammatory diseases, COPD, and obesity in addition to many other known cardiovascular risk factors.

## Introduction

Aortic valve stenosis (AS) has many underlying causes including inflammation, age, and genetic disposition. A study from the National Health, Lung, and Blood Institute demonstrated a prevalence of up to 2.8% in patients over 75 years old, and other studies have shown that its prevalence can reach as high as 9.8% in individuals over 80 years old, with prevalence increasing with age [1]. Once symptomatic, aortic stenosis can carry up to 50% mortality rate [2] and progresses at widely varying rates. Interestingly, the use of statins and the presence of underlying cardiovascular disease risk factors is unrelated to the rate of progression of the condition; only baseline severity has been shown to correlate with the progression rate, with increased baseline severity associated with more rapid AS progression [3]. Aortic stenosis in the elderly occurs due to the degenerative calcification of the aortic cusps. The deterioration of the aortic valve occurs in an active process that shares many features of vascular atherosclerosis. In essence, the aortic valve becomes progressively thickened, fibrotic, and calcified. This occurs by way of a complex mechanism beginning with mechanical stress and endothelial damage and continuing with inflammation, angiogenesis, and hemorrhage in the valve, fibrosis, and, finally, calcification [4].

Previous studies have discussed the epidemiology, molecular mechanisms, and management of AS [1, 4–8] in detail as well as certain cardiovascular risk factors that may contribute to the development of AS [9] without a definitive answer. However, many other risk factors for developing AS are not well known.

Associations between aortic stenosis and many cardiovascular risk factors are strongly suspected [9–12], although most studies on the risk factors of AS contain data from a relative smaller sample size, especially when compared to the data available from the Nationwide Inpatient Sample (NIS) used in this study. The goal of this study is to evaluate any association between aortic stenosis with many comorbidities and race in addition to many common cardiovascular risk factors.

## Methods

### Data Collection and Data Sources

This is a retrospective study using the Nationwide Inpatient Sample (NIS) database. The NIS, developed by the Agency for Healthcare Research and Quality (AHRQ), is a collection of hospital inpatient databases from the Healthcare Cost and Utilization Project (HCUP). HCUP serves as the largest collection of longitudinal hospital care data in the United States. The NIS contains data from millions of hospital stays and covers more than 97% of the US population. The database includes patient demographics, producers, admission, and primary and secondary diagnoses. The data is de-identified, exempt from IRB approval, and publicly available for analysis of nationwide trends in healthcare utilization and outcomes. We analyzed the available database for the years 2016-2020, which contain the first ICD-10 codes separating aortic stenosis from other aortic valve diseases, allowing us to study the risk factors for aortic stenosis alone in order to consider the trends of aortic valve stenosis in patients with a variety of other health conditions including other cardiovascular risk factors, race gender and many inflammatory diseases. . o control for the prevalence of congenital aortic stenosis, we only included patients over the age of 40. However, we did exclude rheumatic valve disease. The International Classification of Disease, Tenth Revision, and Clinical Modification (ICD-10-CM) coding system was used to identify AS patients with accompanying conditions. The ICD-10 code used for aortic stenosis was I35.0. The other codes used in our analysis with their accompanying descriptions are listed Table 1 in the supplementary information. The condition descriptions listed below were taken from the National Center for Health Statistics online database for ICD-10 codes.

**Table 1:** ICD-10 codes with associated conditions used in this study.

| **Condition** | **ICD-10 Code** | **Condition** | **ICD-10 Code** | **Condition** | **ICD-10 Code** |
| --- | --- | --- | --- | --- | --- |
| Dependence on the drug nicotine | F17.200 | Elevated fasting triglycerides | E78.1 | Other rheumatoid arthritis | M06 |
| Dependence on smoking | F19.20 | Hyperchylomicronemia | E78.3 | Scleroderma | M34 |
| Dependence on nicotine chewing tobacco | F17.220 | Kidney disease | N28.9 | Systemic connective tissue disorders | M35 |
| Exposure to environmental tobacco smoke | Z77.22 | Polycystic kidney disease | Q61.3 | Gout | M10 |
| Personal history of tobacco dependence | Z87.891 | Chronic kidney disease stage 1 | N18.1 | Chronic gout | M1A |
| Problem with tobacco use lifestyle | Z72.0 | Chronic kidney disease stage 2 | N18.2 | Psoriatic arthritis | L40.50 |
| Obesity | E66.9 | Chronic kidney disease stage 3 | N18.30 | Cryoglobulinemia | D89.1 |
| Constitutional obesity | E66.8 | Chronic kidney disease stage 3a | N18.31 | Polyarteritis nodosa and related conditions | M30 |
| Morbid obesity | E66.01 | Chronic kidney disease stage 3b | N18.32 | Raynaud's syndrome | I73.0 |
| Essential hypertension | I10 | Chronic kidney disease stage 4 | N18.4 | Buerger's disease (thromboangiitis obliterans) | I73.1 |
| Renovascular hypertension | I15 | Chronic kidney disease stage 5 | N18.5 | Ankylosing spondylitis | M45 |
| Hypertensive heart disease with heart failure | I11.0 | Chronic obstructive pulmonary disease | J44.9 | Antiphospholipid antibody syndrome | D68.61 |
| Hypertensive heart disease without heart failure | I11.9 | Chronic obstructive pulmonary disease with acute exacerbation | J44.1 | Antiphospholipid antibody syndrome with hemorrhagic disorder | D68.312 |
| Hypertensive chronic kidney disease with end stage renal disease | I12.0 | Chronic obstructive pulmonary disease with acute bronchitis | J44.0 |  |  |
| Hypertensive heart and chronic kidney disease with heart failure and stage 1 through 4 chronic kidney disease, or unspecified chronic kidney disease | I13.0 | Bicuspid valve | Q23.1 |  |  |
| Hypertensive urgency | I16.0 | Acquired polycythemia | D75.1 |  |  |
| Dyslipidemia | E78.5 | Benign polycythemia | D75.0 |  |  |
| Mixed hyperlipidemia | E78.2 | Alcohol use | Z72.89 |  |  |
| Other hyperlipidemia | E78.49 | Harmful alcohol use | F10.10 |  |  |
| Elevated cholesterol | E78.00 | Rheumatoid arthritis with rheumatoid factor | M05 |  |  |

### Statistical Analysis

To perform the retrospective univariate and multiple regression analyses on the NIS data from 2016 to 2020 and to adjust for comorbid conditions in the multivariate analysis, patients’ demographic and baseline characteristics will be reported as means, confidence intervals for continuous variables and frequencies, and proportions for categorical variables. Chi-squared/Fisher’s exact test were used to compare categorical variables. For categorical/binary outcomes, multiple logistic regression was used to ascertain the likelihood of the outcome (Odds Ratios (95% CI)) occurring between the characteristics. All statistical models were adjusted for confounding. All analyses were conducted following the implementation of population discharge weights. All p-values were 2-sided and p < 0.05 was considered statistically significant. Data were analyzed using STATA 17 (Stata Corporation, College Station, TX).

## Results

In this study, the NIS data from the years 2016 to 2020 of 112,982,565 patients were analyzed. Of these patients, a total of 2,322,649 were found to have aortic stenosis, with the remaining 110,659,916 patients serving as controls based on their comorbidities. For the years from 2016 to 2020, the data revealed positive associations with several of the studied conditions. Table 2 shows the odds ratios of the univariate analysis. After multivariate analysis, independent positive associations were identified between aortic valve stenosis and the following variables: a bicuspid aortic valve (OR: 11.13, CI 10.38 – 11.93, *P* < 0.001), which was found in 0.63% of patients with AS (14,633 patients) compared to 0.09% of controls (99,594 patients); coronary artery disease (OR: 2.11, CI 2.09 – 2.13, P < 0.001), found in 55.7% of patients with AS (1,293,483 patients) compared to 26.3% of controls (29,103,558 patients); smoking (OR: 1.08, CI 1.07 – 1.08, *P* < 0.001), found in 32.1% of patients with AS (746,499 patients) compared to 26.8% of controls (29,656,857 patients); diabetes mellitus (OR: 1.15, CI 1.14 – 1.16, *P* < 0.001), found in 40.2% of patients with AS (933,937 patients) compared to 32.6% of controls (36,075,133 patients); hypertension (OR: 1.41, CI 1.4 - 1.43, *P* < 0.001), found in 87.7% of patients with AS (2,036,499 patients) compared to 70.4% of controls (77,904,581 patients); hyperlipidemia (OR: 1.31, CI 1.3 – 1.32, *P* < 0.001), found in 62.0% of patients with AS (1,439,113 patients) compared to 42.0% of controls (46,499,297 patients); renal disease (OR: 1.3, CI 1.29 - 1.31, *P* < 0.001), found in 38.8% of patients with AS (900,259 patients) compared to 22.2% of controls (24,522,237 patients); COPD (OR: 1.05, CI 1.04 – 1.05, *P* < 0.001), found in 26.9% of patients with AS (625,722 patients) compared to 20.9% of controls (23,138,988 patients); obesity (OR: 1.3, CI 1.29 – 1.32), found in 9.9% of patients with AS (229,478 patients) compared to 9.2% of controls (10,169,646 patients); rheumatoid arthritis (OR: 1.13, CI 1.11 – 1.15, P < 0.001), found in 2.7% of patients with AS (63,176 patients) compared to 2.4% of controls (2,600,508 patients); scleroderma (OR: 1.93, CI 1.79 – 2.09, P < 0.001), found in 0.18% of patients with AS (4,181 patients) compared to 0.13% of controls (143,858 patients); systemic connective tissue disease (OR: 1.24, CI 1.2 – 1.27, P < 0.001), found in 1.1% of patients with AS (25,317 patients) compared to 0.6% of controls (675,025 patients); polyarteritis nodosa (OR: 1.5, CI 1.24 – 1.81, P < 0.001), found in 0.03% of patients with AS (697 patients) compared to 0.02% of controls (22,132 patients); Raynaud’s syndrome (OR: 1.16, CI 1.09 – 1.24, P < 0.001), found in 0.26% of patients with AS (6,039 patients) compared to 0.22% of controls (243,452 patients); Caucasian race (OR: 1.47, CI 1.42 – 1.52, *P* < 0.001), a characteristic of 83.4% of patients with AS (1,937,322 patients) compared to 71.0% of controls (78,601,738 patients). There was a multivariate odds ratio of 1.08 for each additional year of age (P < 0.001) and male gender (OR: 1.21, CI 1.2-1.23 *P* < 0.001), Figures 1, 2, Tables 1 and 2]. Other ethnicities included in the analysis, including African American, Hispanic, Asian/Pacific Islander, and Native American, exhibited decreased odds ratios of AS. The odds ratio was 0.55 (CI 0.54 – 0.56, P < 0.001) for African American with 6.5% of patients with AS compared to 14.3% of controls, 0.75 (CI 0.74 – 0.77, P < 0.001) for Hispanic with 6% of patients with AS compared to 9.1% of controls, 0.62 (CI 0.59 – 0.64) for Asian/Pacific Islander with 1.7% of patients with AS compared to 2.3% of controls, and 0.71 (CI 0.67 – 0.75, P < 0.001) for Native American with 0.3% of patients with AS compared to 0.6% of controls. No meaningful associations were identified between AS and polycythemia, alcohol use, gout, psoriatic arthritis, Buerger’s disease, ankylosing spondylitis, and antiphospholipid antibody syndrome, There appears to be a linear relationship between age and development of aortic stenosis (figure 3).

**Figure 1:**
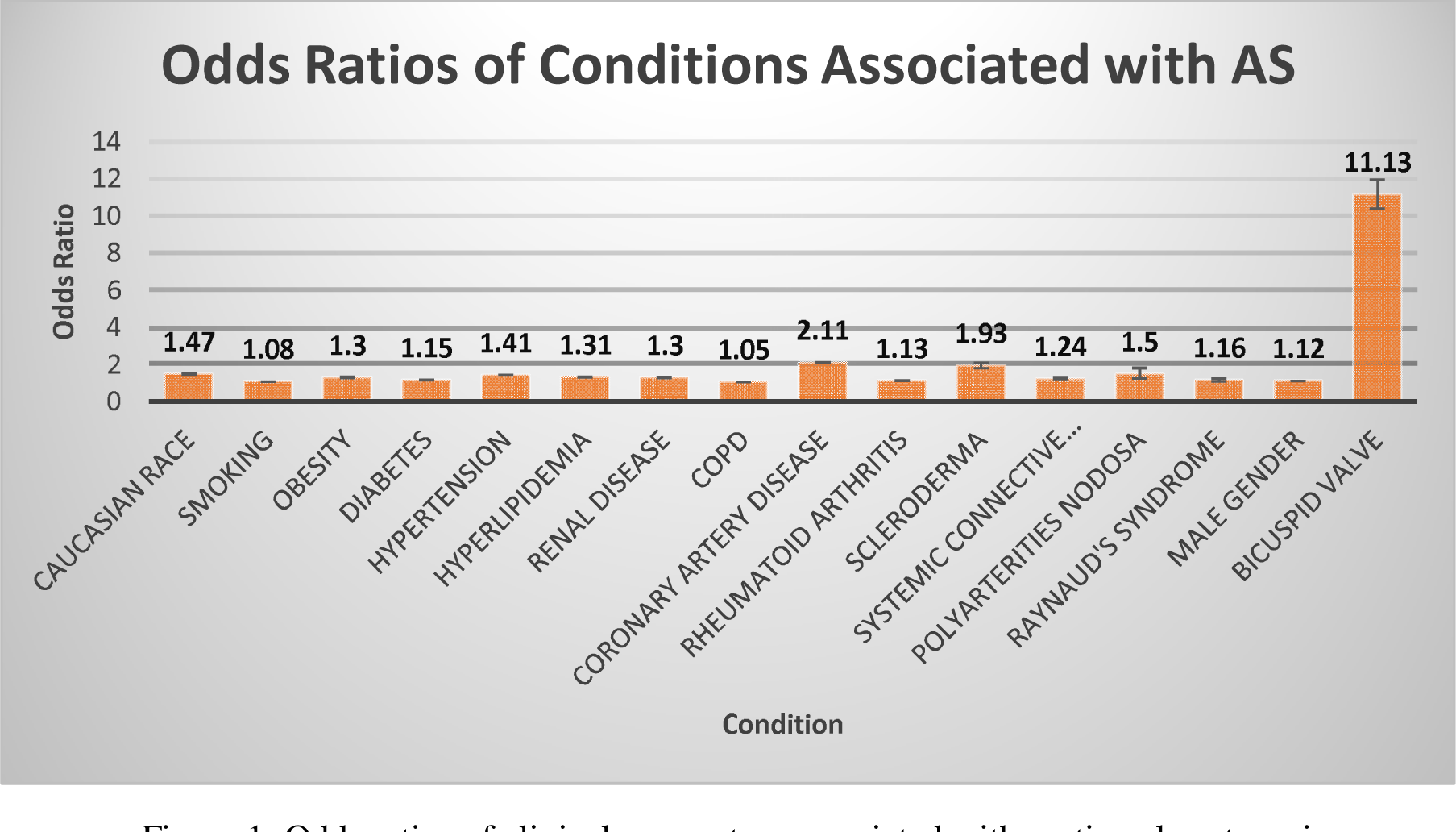
Odds ratios of clinical parameters associated with aortic valve stenosis

**Figure 2:**
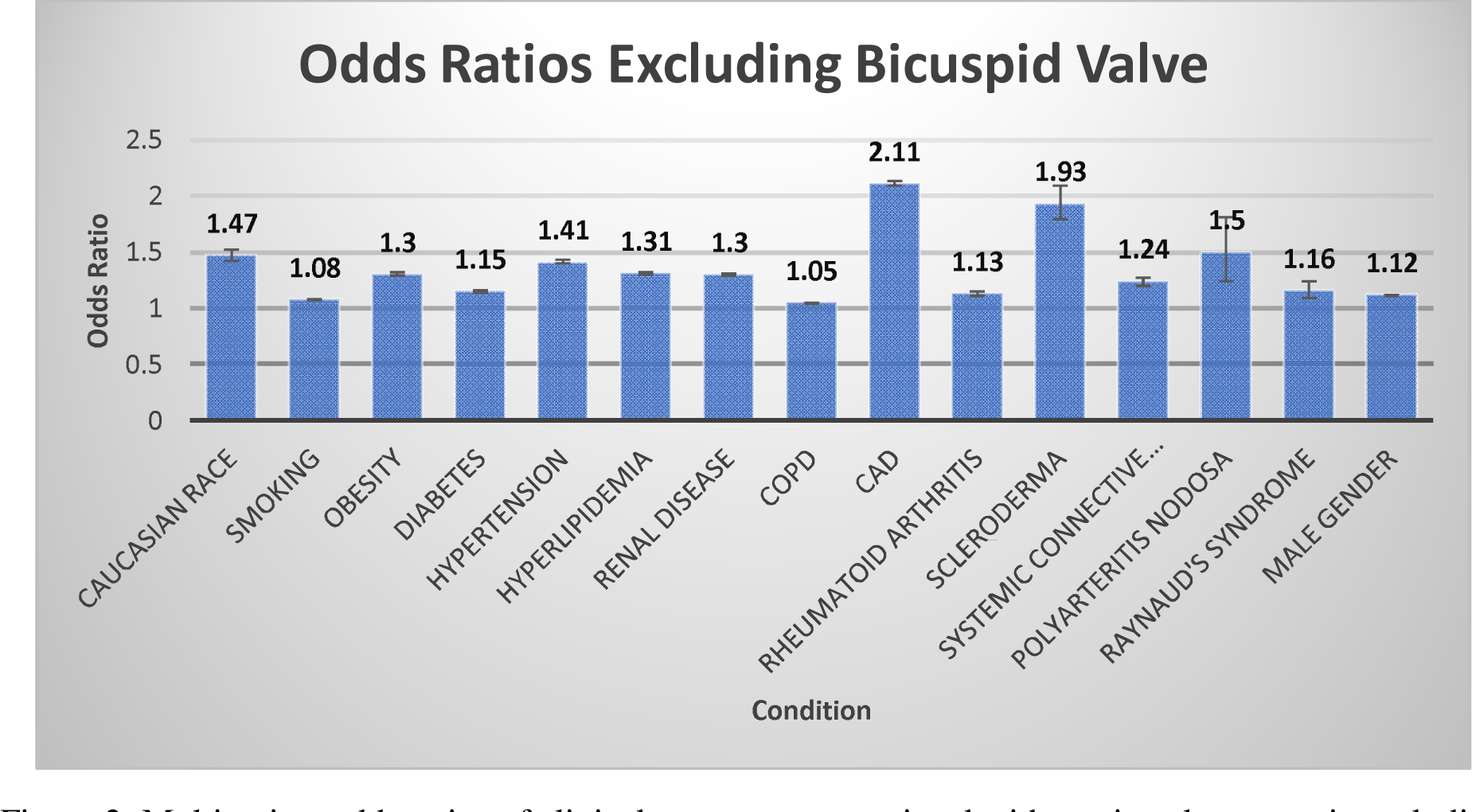
Multivariate odds ratios of clinical parameters associated with aortic valve stenosis excluding commonly known causes such as bicuspid aortic valve

**Figure 3:**
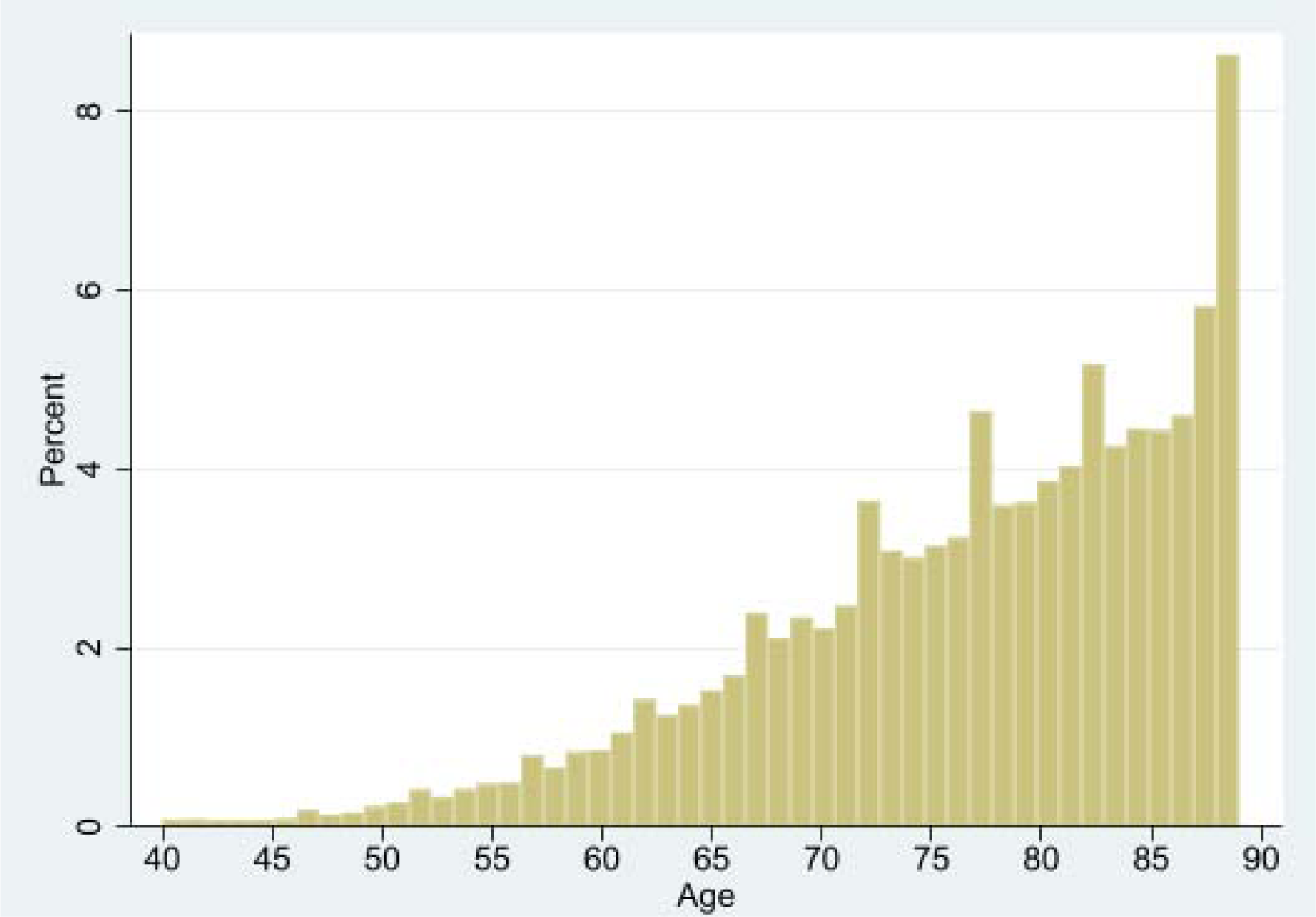
A linear increase in the prevalence of aortic stenosis with aging.

**Table 2:**
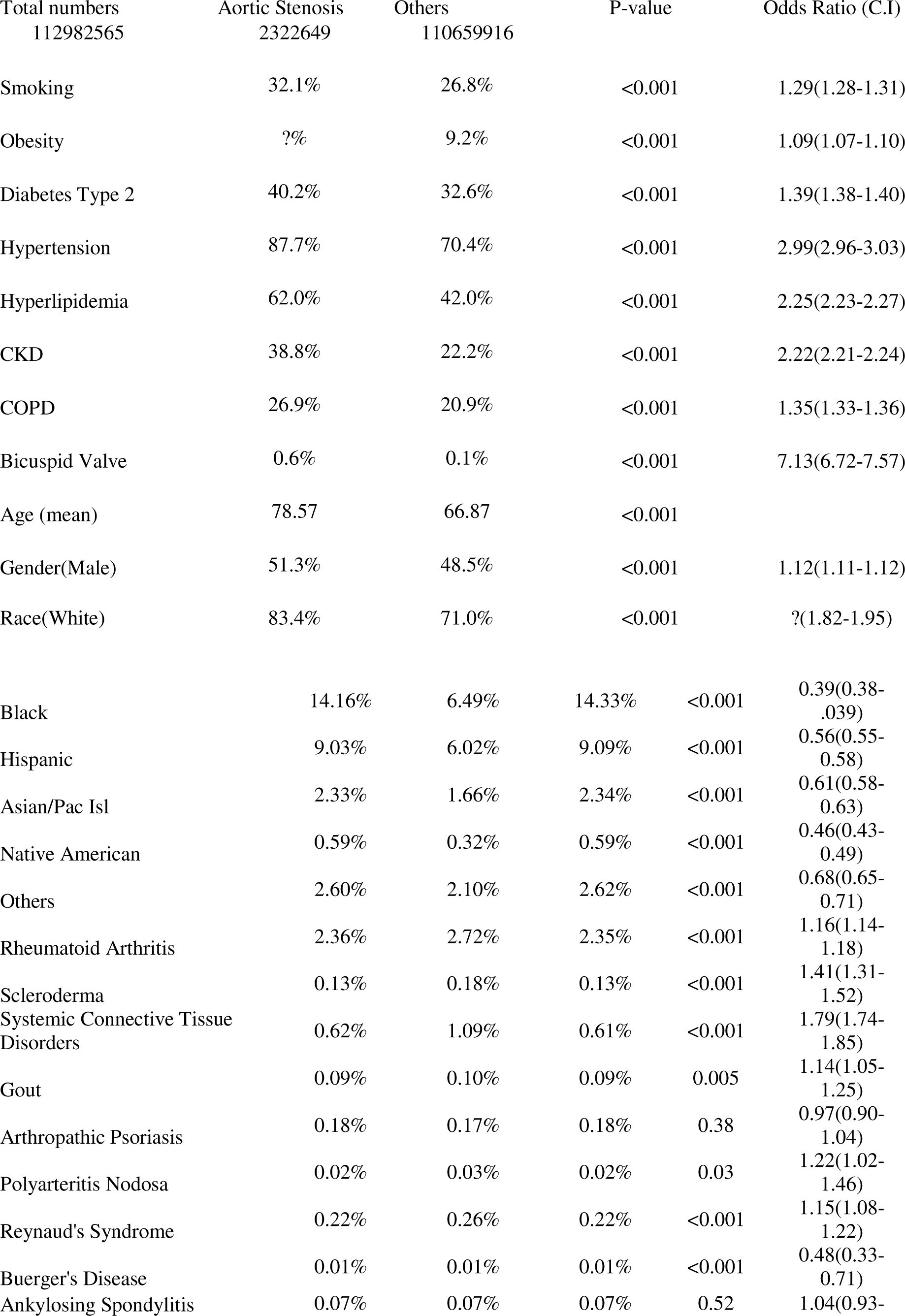
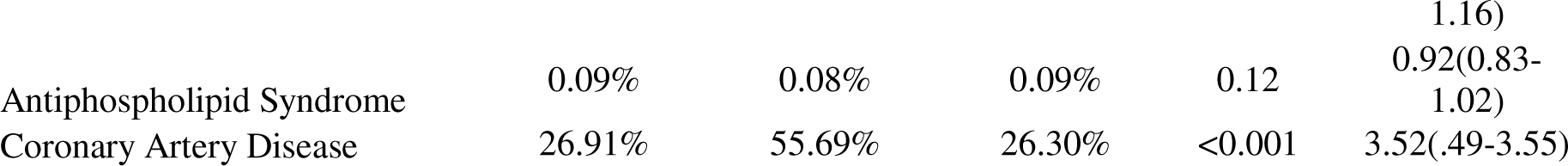
Univariate data for aortic stenosis association with comorbid conditions, age and race.

**Table 3:** Multivariate odds ratios of parameters studied with confidence intervals.

| Condition | Odds Ratio | Confidence Interval | P-value |
| --- | --- | --- | --- |
| Age for each year | 1.07 | 1.07 – 1.07 | < 0.001 |
| Caucasian race | 1.47 | 1.42 - 1.52 | < 0.001 |
| Smoking | 1.08 | 1.07 - 1.08 | < 0.001 |
| Obesity | 1.3 | 1.29 - 1.32 | < 0.001 |
| Diabetes mellitus | 1.15 | 1.14 - 1.16 | < 0.001 |
| Hypertension | 1.41 | 1.4 - 1.42 | < 0.001 |
| Hyperlipidemia | 1.31 | 1.3 - 1.32 | < 0.001 |
| Renal disease | 1.3 | 1.29 - 1.31 | < 0.001 |
| COPD | 1.05 | 1.04 - 1.05 | < 0.001 |
| Male gender | 1.12 | 1.11 – 1.12 | < 0.001 |
| Rheumatoid arthritis | 1.13 | 1.11 – 1.15 | < 0.001 |
| Scleroderma | 1.93 | 1.79 – 2.09 | < 0.001 |
| Systemic connective tissue disorders | 1.24 | 1.2 – 1.27 | < 0.001 |
| Polyarteritis nodosa | 1.5 | 1.24 – 1.81 | < 0.001 |
| Raynaud's syndrome | 1.16 | 1.09 – 1.24 | < 0.001 |
| Bicuspid valve | 11.13 | 10.38– 11.93 | < 0.001 |

## Discussion

Aortic valve stenosis is a very common heart valve disease, especially among elderly patients. The results of this study demonstrated significant correlations between aortic valve stenosis and a variety of cardiovascular and other comorbidities including age and race. Each of these conditions will be discussed in more detail in the following paragraphs.

There are three risk factors that have a particularly well-documented association with AS: a congenital bicuspid aortic valve, rheumatic heart disease, and advanced age. Rheumatic heart disease was excluded from this study. A bicuspid aortic valve is the most common congenital heart valve disease, with a prevalence of up to 2% [2]. Consistent with the literature on the subject, this retrospective study found that those with a bicuspid aortic valve had over 11 times higher odds of developing AS than those without. Age has been widely associated with the development of AS. This study found more than 8 times higher odds of AS in those aged above 60 than those of younger ages but also found that each year of aging increased risk of aortic stenosis. This association of age with AS occurs by way of several mechanisms [2].

Smoking has long been identified as a risk factor for the development of cardiovascular disease [7], and this study serves as another confirmation of the positive association between aortic stenosis and smoking. The association of aortic stenosis and smoking in combination with the association of AS. The Li group hypothesized that smoking, which introduces injurious chemicals into the cardiovascular system, may worsen an already faulty valve repair system in certain patients which eventually results in excessive valve thickening [7]. A Japanese study identified similar results [13]. These findings, along with those of this study, highlight the importance of smoking as a modifiable risk factor in the development of AS.

Obesity is another condition that has been well-established as a significant risk factor for the development of cardiovascular disease and the incidence of cardiovascular events. A strong correlation between obesity and aortic valve disease was identified in this study. Interestingly, the literature is somewhat divided on the relationship between obesity and aortic stenosis. The findings of this study add to the controversy surrounding the role of obesity in the development of AS, with some retrospective studies finding positive correlations [14, 15] and others finding no significant association [16]. In fact, some studies have shown lower rates of ischemic cardiovascular events and AS-related events in those with an overweight body mass index (BMI) in what has been termed the “obesity paradox” [17]. One possible explanation for this is that obese individuals may also receive more aggressive treatment for obesity-induced comorbidities [18], which may lead to earlier intervention that partly prevents processes that lead to AS. The strong positive correlation identified in this study, however, falls in line with more recent retrospective studies that have shown stronger associations between AS and obesity [19]. In addition, in 2020, Kaltoft et al. established a genetic association between obesity and the risk of developing AS, hypothesizing the cause to be a combination of structural and metabolic abnormalities present in obese patients [20]. There are many proposed associations and mechanisms for the increase in risk of AS associated with obesity, including changes in the distribution of adiposity, changes to the sympathetic nervous system, sympathetic dysregulation, direct effects on cardiac myocytes, changes to the cardiac natriuretic peptide system, inflammation, hemodynamic disturbance, and others [21].

Diabetes mellitus has been identified as playing a role in the development of cardiovascular disease since a link between the two was uncovered by the Framingham Heart Study in the late 1970s. In particular, diabetes has been associated with aortic stenosis [22], and this study confirms this association with diabetic patients exhibiting a 15% greater odds ratio when compared to controls. The diabetic state appears to accelerate most cardiac pathologies via a variety of mechanisms, including causing abnormalities in systemic and local vascular inflammation, endothelial and microvascular injury, altered thrombosis, autonomic nerve dysfunction, and membrane instability in nerves, smooth muscles, and endothelium [23].

Another condition that has been strongly associated with cardiovascular disease is hypertension. The condition affects up to a third of the adult population of the United States. Hypertension has also been associated with AS [24], and this study confirms this association, with a 41% higher odds ratio of AS in patients with hypertension. The increase in systemic vascular resistance that can occur by way of a variety of mechanisms causes anomalous intracardiac pressures in the left heart, leading to left ventricle damage. [25]. A 2013 study found that the progression of calcific aortic valve disease was faster in hypertensive patients and slower in patients taking angiotensin-receptor blockers [26].

Hyperlipidemia is a common condition that clinicians strive to quickly address due to its central role in atherosclerosis and its strong association with many cardiovascular diseases. Various forms of dyslipidemia are associated with atherosclerosis due to the various effects of low-density lipoprotein (LDL) on arteries. In fact, previous studies have shown that atherosclerosis risk factors and proximal aortic atherosclerosis are independently associated with aortic valve abnormalities, specifically aortic valve sclerosis [14]. This study supports these findings with the identification of a 31% higher odds ratio of AS in hyperlipidemic patients. The development of AS has been demonstrated to occur at early stages in patients with homozygous familial hypercholesterolemia, a condition in which patients have very high levels of LDL-C [27].

Coronary artery disease is a common condition that is one of the ultimate manifestations of many of the risk factors discussed in this work. This study found an odds ratio of 2.11 for the development of AS in patients with coronary artery disease. Coronary artery disease has been extensively studied in its relationship with many conditions, including diabetes, hypertension, hyperlipidemia, and others. It is important to note that the odds ratio found in this study signifying a greater risk of AS in patients with coronary artery disease is likely the result of many AS patients demonstrating several of the well-established cardiovascular disease risk factors mentioned previously and does not necessarily mean or suggest that coronary artery disease directly causes AS. Aortic stenosis may be more strongly related to the cardiovascular disease risk factors such as hypertension, hyperlipidemia, etc., which also each individually contribute to the development of coronary artery disease. Therefore, AS and coronary artery disease may simply be two co-existing conditions that result from these underlying disorders, but more likely there is a complex interplay between the two that results in development and progression of both. Without further study, however, it is difficult to more precisely identify and characterize the relationship between coronary artery disease and AS.

Chronic kidney disease has been associated with increased cardiovascular morbidity and mortality. In fact, even the early stages of chronic kidney disease have been associated with increased calcification of the heart valves and coronary arteries [28]. The aortic valve has also been shown to be particularly affected by this calcification, with dialysis patients having a higher rate of progression of AS [29]. The present large-database study found a 30% higher odds ratio of AS in those with documented renal disease than in those without. The advanced progression of AS in renal disease is thought to occur mainly due to irregularities in calcium homeostasis, including increased parathyroid hormone, calcium-phosphate products, and excess vitamin D [30].

The association of aortic stenosis with chronic obstructive pulmonary disease (COPD) has not been well-explored. Most publications focus on the risk of aortic valve replacement in patients with COPD, and it has been noted that the difficulty of determining the precise contribution of each pathology may be due to some of the overlapping symptoms, such as progressive dyspnea [31]. Even after adjusting for smoking, the present study found a significant positive association between aortic stenosis and COPD, with an odds ratio 5% higher in those with COPD compared to controls. Similar to certain other risk factors previously discussed, COPD may contribute to the development of AS through direct and indirect mechanisms. One way is through its contribution to atherosclerosis, which has been well-documented [32]. The chronic inflammation present in COPD may also contribute to sustained levels of transforming growth factor-β (TGF-β), a peptide that stimulates the formation of extracellular matrix and that is found at higher levels in states of repeated tissue injury. TGF-β has been suggested to play a significant role in the pathogenesis of AS [8]. A more direct relationship between COPD and AS has been suggested by one small Turkish study that found that patients with COPD exhibited enhanced calcification within stenotic lesions compared to controls [33]. As previously discussed, calcification is a major causative factor of AS, and these results combined with those of this study are suggestive of the role that COPD plays in the development of AS. The exact mechanisms by which this occurs, however, are yet to be understood.

As previously acknowledged, chronic inflammation has been strongly suspected in the pathogenesis of aortic stenosis. It is therefore reasonable to expect that conditions that introduce widespread inflammation in the body may affect the integrity of the aortic valve. Isolated cases of patients with both AS and autoimmune disease have been reported, but no large studies have been performed to detect associations between AS and certain autoimmune diseases. In addition, cardiovascular involvement is a well-known complication of several autoimmune diseases, including rheumatoid arthritis and scleroderma, but there are relatively few studies that report valvular disease in such cases. Interestingly, significant positive associations were found in the present study between AS and several of the autoimmune diseases included, but not all of them. In particular, positive associations were found between AS and rheumatoid arthritis, scleroderma, systemic connective tissue disease, polyarteritis nodosa, and Raynaud’s syndrome. No meaningful associations were found between AS and psoriatic arthritis, Buerger’s disease, gout, ankylosing spondylitis, or antiphospholipid antibody syndrome. Some of the inflammatory conditions in this study have been suggested to have associations with AS in very small studies or case reports, including rheumatoid arthritis [34], mixed connective tissue disease [35], and scleroderma (systemic sclerosis) [36] . For others, such as polyarteritis nodosa and Raynaud’s syndrome, this is the first known identification of a relationship. Without further study, it is difficult to determine why only some of the inflammatory syndromes were found to be associated with AS while others were not. Therefore, the mechanisms whereby certain inflammatory syndromes may lead to aortic valve changes while others do not warrant further investigation and exploration.

Finally, this study found a significantly higher odds ratio of AS of up to ?% in Caucasian patients compared to other races recorded in the database, including African American, Hispanic, Asian/Pacific Islander, and Native American. The non-Caucasian ethnicities were each found to have significantly reduced odds of developing AS (see Results). This finding has been validated by other studies that include other non-white ethnicities [37] and is in agreement with other studies in the literature that have found an increased prevalence of AS in Caucasian patients, including one large study that analyzed a database of over 2.1 million de-identified patients [38]. In particular, the reduced odds ratios of other races compared to Caucasians in this study confirm those in a recent study that also used the NIS to estimate odds ratios of AS based on race [39]. Other studies have noted that African American patients have significantly less AS, aortic valve calcification, degenerative valve disease, and bicuspid aortic valves than Caucasian patients [40]. It is possible that the finding in this study is due to the overwhelming Caucasian representation in the NIS database, with Caucasian patients comprising more than 71% of the included individuals. African Americans, Hispanics, Asian/Pacific Islanders, Native Americans, and other races comprised 14.2%, 9%, 2.3%, 0.6%, and 2.6% of the database, respectively. Possible explanations for these findings include differing socioeconomic status and relatively later presentation to healthcare professionals. The precise reasons for the much lower incidence of AS in African Americans and other ethnicities are unclear, warranting further investigation.

## Limitations

The results of this study are limited by its retrospective nature. Coding inaccuracies in the ICD-10 system may have introduced errors into our analysis as well. The sheer number of coded conditions and subdivisions in the ICD-10 system made it difficult to include each and every relevant condition from each of the categories that were studied, but it is the opinion of the authors that the main contributors for each condition were included in this study. The choice of which specific ICD-10 codes to include in each disease category may have also introduced error in the detected associations between AS and the diseases studied. In addition, because the NIS is strictly an inpatient sample database, our results do not include those who may be outpatients and thus may not be an accurate reflection of outpatient data.

## Conclusions

Using a large inpatient database from the years 2016 to 2020, we found strong positive associations between aortic stenosis and known cardiovascular risk factors such as age, bicuspid aortic valve, smoking, diabetes mellitus, hypertension and hyperlipidemia. Furthermore, we found strong independent associations between aortic stenosis and renal disease, COPD, polyarteritis nodosa, Raynaud’s syndrome, systemic connective tissue disease, rheumatoid arthritis, scleroderma, and Caucasian race. No meaningful associations were found between aortic stenosis and alcohol use, polycythemia vera, gout, Buerger’s disease, or systemic lupus erythematosus. The mechanisms behind each of these associations are very complex, warranting further investigations.

## Data Availability

Yes

https://www.ahrq.gov/data/hcup/index.html

## Supplementary Information

